# Analysis of uptake, effectiveness and safety of COVID-19 vaccinations in pregnancy using the QResearch® database: research protocol and statistical analysis plan

**DOI:** 10.1101/2022.12.19.22283660

**Authors:** Emma Copland, Jennifer Hirst, Tom Ranger, Winnie Mei, Sharon Dixon, Carol Coupland, Kenneth Hodson, Jonathan Luke Richardson, Anthony Harnden, Aziz Sheikh, Carol Dezateux, Brenda Kelly, Marian Knight, Johnathan van Tam, Alessandra Morelli, Joanne Enstone, Julia Hippisley-Cox

## Abstract

**Background:** The COVID-19 pandemic has affected millions of people globally with major health, social and economic consequences, prompting development of vaccines for use in the general population. However, vaccination uptake is lower in some groups, including in pregnant women, because of concerns regarding vaccine safety. There is evidence of increased risk of adverse pregnancy and neonatal outcomes associated with SARS-CoV-2 infection, but fear of vaccine-associated adverse events on the baby both in short and longer term is one of the main drivers of low uptake for this group. Other vaccines commonly used in pregnancy include influenza and pertussis. These both have reportedly higher uptake compared with COVID-19 vaccination, which may be because they are perceived to be safer. In this study, we will undertake an independent evaluation of the uptake, effectiveness and safety of COVID-19 vaccinations in pregnant women using the QResearch primary care database in England.

**Objectives:**

A. To determine COVID-19 vaccine uptake in pregnant women compared to uptake of influenza and pertussis vaccinations.
B. To estimate COVID-19 vaccine effectiveness in pregnant women by evaluating the risk of severe COVID-19 outcomes following vaccination.
C. To assess the safety of COVID-19 vaccination in pregnancy by evaluating the risks of adverse pregnancy and perinatal outcomes and adverse events of special interest for vaccine safety after COVID-19 vaccination compared with influenza and pertussis vaccinations.

**Methods:** This population-based study uses the QResearch® database of primary health care records, linked to individual-level data on hospital admissions, mortality, COVID-19 vaccination, SARS-CoV-2 testing data and congenital anomalies. We will include women aged 16 to 49 years with at least one pregnancy during the study period of 30^th^ December 2020 to the latest date available. Babies born during the study period will be identified and linked to the mother’s record, where possible.

We will describe vaccine uptake in pregnant women by trimester and population subgroups defined by demographics and other characteristics. Cox proportional hazards multivariable regression will be used to identify factors associated with vaccine uptake. The effectiveness of COVID-19 vaccines in pregnant women will be assessed using a nested matched case-control design to assess hospitalisation, intensive care admission and death with COVID-19. Cases who had the outcome will be matched with up to 10 controls who did not have the outcome on that date by age, calendar date and trimester of pregnancy using incidence density sampling for the occurrence of each outcome after each vaccine dose compared with unvaccinated individuals. For the safety analysis, we will we use logistic regression analyses to determine unadjusted and adjusted odds ratios for the occurrence of maternal (e.g. miscarriage, ectopic pregnancy and gestational diabetes) and perinatal outcomes (e.g. stillbirth, small for gestational age and congenital anomalies) by vaccination status compared to unvaccinated individuals. For the adverse events of special interest for vaccine safety (e.g. venous thromboembolism, myocarditis and Guillain Barre syndrome), we will use time varying Royston-Palmar regression analyses to determine unadjusted and adjusted hazard ratios for the occurrence of each outcome by vaccination status to unvaccinated individuals.

**Ethics and dissemination:** QResearch is a Research Ethics Approved Research Database with ongoing approval from the East Midlands Multi-Centre Research Ethics Committee (Ref: 18/EM/0400). This study was approved by the QResearch Scientific Committee on 9^th^ June 2022. This research protocol has been developed with support from a patient and public involvement panel, who will continue to provide input throughout the duration of the study. Research findings will be submitted to pre-print servers such as MedRxIv, academic publication and disseminated more broadly through media releases and community groups and conference presentations.

## Background

On 11th March 2020, the World Health Organization (WHO) declared a global COVID-19 pandemic, which has since affected millions of people globally with major health, social and economic consequences. This prompted the rapid development and licensing of several vaccines for use in the general population. Currently in Great Britain everyone 12 years of age and older is eligible to get a COVID-19 vaccination. The COVID-19 vaccines currently approved for use in the UK are Pfizer/BioNTech, Moderna, Oxford/AstraZeneca, Nuvaxovid vaccine (Novavax), Janssen and Valneva, although the only three currently in use are Pfizer/BioNTech, Moderna and Nuvaxovid vaccine (Novavax). The Oxford/AstraZeneca vaccine has been withdrawn from use in the UK since August 2022.(1)

As of September 2022, 62% of the global population has currently been fully vaccinated,(2) and 70% of people in England have received at least one dose.(3) Initial advice on vaccination in pregnancy was only for those in higher risk groups or who were clinically vulnerable due to lack of data in pregnant women. Due to the lack of information on the safety profile pregnant women were excluded from the initial clinical guidance. In April 2021, the advice on vaccination was extended to include all pregnant women. Yet, even after recommendation, concerns amongst pregnant women regarding vaccine safety have been a barrier to vaccine acceptance. There is clear evidence from observational data that pregnant women are at increased risk of severe illness from COVID-19 compared with non-pregnant women, particularly in the third trimester (4) and these are higher in people living with comorbidities and those of non-white ethnicity.(5) Furthermore, having COVID-19 during pregnancy is also associated with higher risk of adverse baby outcomes, including stillbirth and neonatal death.(5) It is known that the risk of hospitalisation, intensive care unit admission, maternal death, baby deaths and stillbirth are vastly reduced following vaccination in pregnancy.(6–10) However, more data are needed on rare outcomes to help to inform operational decisions about the use and distribution of vaccines and to obtain fully informed consent for vaccination.

As of 16 April 2021, the Joint Committee on Vaccination and Immunisation (JCVI) advised that pregnant women should be offered COVID-19 vaccines at the same time as people of the same age or risk group.(11, 12) However, in May 2021, only 2.8% of women giving birth had received at least one dose of vaccine. This increased to 22.7% in August 2021, 53.8% in December 2021 and 73.2% in May 2022. As of May 2022, 26.5% of women were unvaccinated at time of delivery. Vaccination is known to be lower in younger women, people living in areas of high deprivation and in Black, Asian and Afro-Caribbean ethnic groups.(13) In the overall 17-month period between January 2021 and May 2022 a total of 717,977 women gave birth in England of whom 231,082 (32.3%) had received at least one dose of COVID-19 vaccine prior to delivery.(14, 15) Within women who were vaccinated during pregnancy, 55,729 received at least one dose in the first trimester, 83,585 in the second trimester and 70,972 in the third trimester. In December 2021, pregnant women were added to the UK’s priority vaccine list and will be prioritised for the Autumn 2022 booster dose.(16)

COVID-19 vaccines are recommended for pregnant women to prevent severe consequences of COVID-19 in pregnancy, including admission of the woman to intensive care and adverse birth outcomes such as stillbirth and premature delivery.(17) A population-based cohort study in England (18) of 342,080 women has shown that SARS-CoV-2 infection at the time of birth is associated with higher rates of fetal death, preterm birth, preeclampsia, and emergency caesarean birth. Furthermore, a study using population-level data in Scotland has revealed that 98% of critical care admissions due to SARS-CoV-2 in pregnant women and all new-born baby deaths occurred in women who were unvaccinated at the time of COVID-19 diagnosis.(6, 19) A review of 26 studies found no significant increase in the rates of preterm birth, fetal growth restriction, caesarean birth and neonatal intensive care unit admission after vaccination in pregnant patients compared with those who were unvaccinated.(20) This review also identified 12 studies evaluating vaccine adverse effects in pregnancy which suggested that the vaccine side effect profile is similar to that in non-pregnant people.

Despite the evidence of increased risk of adverse pregnancy and neonatal outcomes associated with SARS-COV-2 infection in pregnancy, the fear of vaccine-associated adverse events on the baby both in short and longer term is one of the main drivers to low vaccine uptake for this group. A study including 35,691 pregnant women aged 16 to 54 years old in the USA (21) using the spontaneous adverse events reporting data has not shown any obvious safety signals among pregnant women who received the Pfizer/BioNTech and Moderna vaccines.

In this context, understanding how the risks of adverse pregnancy outcomes compare between those vaccinated in pregnancy and those who were not would provide evidence of the likely impact of COVID-19 vaccination on pregnancy outcomes. This robust evidence can then be made available to clinicians, policy makers and the public so that it can be used at the point of care to ensure that pregnant women can make informed decisions as to whether to have the vaccine. Further information on how the risk of the same adverse pregnancy outcomes compare following a SARS-CoV-2 infection during pregnancy is needed.

Vaccination uptake has been reported to be lower in pregnant women from the most disadvantaged parts of the UK and in ethnic minorities.(14, 22) Uptake in the most disadvantaged and in ethnic minority groups tends to be much lower until high levels of coverage have been achieved in the wealthier groups.(23) Therefore, the mass COVID-19 vaccination programme could not only exacerbate existing health inequalities regarding uptake of the vaccination but could also compound inequalities for disadvantaged and ethnic minority groups who are already known to be at higher risk of severe outcomes from COVID-19 infection.(24)

Other vaccines that are commonly administered in pregnancy in primary care are influenza and pertussis.(25) These have been recommended for pregnant women for a number of years and, although uptake is better than COVID-19 vaccination uptake, it still remains relatively low at 45% for influenza,(25) lower than the national average. It is likely that the low vaccination rates reflect general safety concerns, and result in potentially preventable maternal morbidity and mortality. Pertussis vaccination in pregnancy was rolled out in the UK in 2012 in response to a national outbreak to protect infants in the first months of life and is ideally given between weeks 20 and 32 of pregnancy. Uptake is higher than influenza, reported as 64.5% in spring 2021.(26) Evidence suggests that uptake of COVID-19 vaccines is lower than either influenza or pertussis, which may be because women are concerned about and the shorter- and longer-term safety for themselves and their babies.

In addition to vaccine safety and effectiveness monitoring in pregnancy, it is important to monitor vaccine safety outcomes by trimester of pregnancy, as there is currently only limited information on whether outcomes differ by trimester of vaccination including pre-conception.(21, 27) Furthermore, stratification by socioeconomic position, educational level, and ethnic group will help healthcare professionals and policy makers understand which groups are least likely to be vaccinated and who is at highest risk of adverse outcomes.

The Pfizer/BioNTech or Moderna vaccines are the preferred vaccines for use during pregnancy,(28) but these recommendations may only have arisen because the first vaccine safety reports in pregnancy used Pfizer/BioNTech or Moderna which were the first vaccines available in the US.(21) There is scant evidence to suggest that one vaccine may be safer than another, so it is important to understand whether there are differences in efficacy and safety between the different COVID-19 vaccinations during pregnancy.

In this study, we will undertake an independent evaluation of the uptake, effectiveness and safety of COVID-19 vaccination in pregnant women during the COVID-19 pandemic using the QResearch primary care database linked to mortality, hospitalisation, infection, maternity service and health outcome information. This work will build on a series of studies on COVID-19 vaccination safety and effectiveness published in leading journals (29–31) (including the British Medical Journal and Nature Medicine), which generated considerable policy, scientific, and public interest and provided key information for policy makers.

The QResearch dataset covers approximately 20% of UK primary care, representing 13 million patients across the UK. Through linkages with vaccination, maternity service data and health outcome datasets, we will be able to evaluate and compare geographic differences in uptake and safety and assess rare outcomes including congenital anomalies and to compare COVID-19 vaccination with other vaccinations commonly administered in pregnancy. We will also evaluate how the risk of safety outcomes associated with vaccination in pregnancy compares with the risk of the same outcomes after contracting SARS-CoV-2 infection in pregnancy. A strength of this study is that it will set up data and methodology for investigating post-marketing vaccine safety and effectiveness in pregnant women.

## Objectives

### Objective A: Uptake of vaccines in pregnant women

We will determine COVID-19 vaccine uptake in pregnant women by vaccine type and number of doses overall. We will also evaluate:

A1: Uptake of COVID-19 vaccines in pregnant women over time and by trimester and in subgroups including age group, ethnicity, deprivation, geographic region of the UK, co-morbidities, QCovid risk score and prior COVID-19 status.

A2: Uptake of influenza and pertussis overall and stratified by socio-demographic factors (as detailed above).

### Objective B: Effectiveness of COVID-19 vaccines in pregnant women

We will estimate vaccine effectiveness in pregnant women by evaluating the risk of a severe COVID-19 outcome (hospital or intensive care unit (ICU) admission or death) following one, two or three or more doses of vaccination, by vaccine type and virus strain, where numbers are sufficient. We will also evaluate:

B1. effectiveness of COVID-19 vaccines against severe COVID-19 outcomes in pregnant women in subgroups (where numbers permit) including age group, ethnicity, deprivation, co-morbidities, trimester of vaccination, and QCovid risk score and prior COVID-19 status;

B2. the risk of a severe COVID-19 diagnosis following a SARS-CoV-2 infection during pregnancy or a combination of SARS-CoV-2 infection and vaccination.

### Objective C: Safety of vaccines in pregnant women

We will evaluate the risks of pregnancy loss, adverse neonatal outcomes and other safety outcomes in pregnant women following COVID-19 vaccination (such as thrombosis) by vaccine type and dose and the risk of relevant adverse events of special interest for vaccination. In addition, we will evaluate:

C1. safety of COVID-19 vaccines in pregnant women by trimester and where numbers are sufficient, within subgroups including by age group, ethnicity, deprivation, co-morbidities, QCovid risk score and prior COVID-19 status by comparing the risks of adverse outcomes in those who were vaccinated and those who were not;

C2. how the risks of these adverse outcomes associated with vaccination in pregnancy compares with the risks associated with SARS-CoV-2 infection in pregnancy.

C3. Safety of influenza and pertussis vaccines overall and stratified by socio-demographic factors (as detailed above).

## Methods

### Study period

Our study period is from 30^th^ Dec 2020, when pregnant women first became eligible for COVID-19 vaccination to the latest date for which data are available at the time of the analysis.

### Inclusion criteria

We will identify pregnant women during the study period aged 16-49 years registered with participating primary care practices in QResearch. This will be achieved using established algorithms for identifying pregnancies from the GP record (32) as well as utilising the Hospital Episodes Statistics (HES) linked datasets. Babies born on or after 30^th^ December 2020 will also be included in the study. We will link the records of these pregnant women with the records of their babies where possible.

### Exclusion criteria

We will exclude temporary residents, people without a pregnancy in the study period and babies born before the 30^th^ December 2020.

### Data sources and settings

Our main analyses will be based on the QResearch GP database linked to the following NHS Digital datasets to improve ascertainment of outcomes, exposures and confounders:

#### Existing data linkages which are updated monthly

- QResearch database 1500 general practices in England, covering a current population of 13 million patients. This includes demographics, diagnoses, medication, laboratory investigations, pregnancy information, referrals. It is estimated that there will have been over 700,000 pregnancies in England since the start of the COVID-19 vaccine rollout.(33)
- SGSS (Second Generation Surveillance System) data which includes Pillar 1 and Pillar 2 SARS-CoV-2 testing data which includes individual PCR test results from hospital and community settings for positive results with the date and S-gene drop out (used as a proxy for variant type).
- Civil Registration Data which includes individual level data for date and cause of death
- HES care data which includes individual level data for hospital admissions, critical care, outpatients and A&E attendances
- SUS-PLUS (Secondary Uses Service) data which is similar to HES data but which is available with 2-3 weeks latency
- COVID-19 Vaccine uptake data from the National Immunisation Database which includes vaccine type, date, dose
- COVID-19 vaccination adverse events dataset within 15 minutes of administration

#### New data linkages

We will undertake two new data linkages which will improve ascertainment of key outcomes

- Congenital anomalies registry (NCARDRS)
- Maternity Services Data Set (MSDS)

In addition, we will compare results and validate signals in Scottish data sets led by Professor Aziz Sheikh.(6, 34)

### Study Design

This is a series of observational studies using routinely linked electronic records. We will utilise two main study designs: a cohort study and nested case-control studies. Prior to undertaking the statistical analysis, we will define the pregnant cohort by estimating the date of conception and duration of each pregnancy, and the start and end dates for each trimester using the following methodology:

### Determining pregnancy outcomes to derive pregnancy end date

We will identify the first pregnancy outcome in each woman’s record using linked GP and HES records and the date on which this outcome occurred. Pregnancy outcomes consist of delivery (live/still birth; single or multiple birth; full, pre-term or post-term), ectopic pregnancy, termination of pregnancy, miscarriage, molar pregnancy, other codes linked to early pregnancy loss, blighted ovum and codes linked to postnatal care.

We will identify subsequent outcomes to determine multiple pregnancies for individuals during the study period. We will consider delivery outcomes that occur at least 25 weeks after a prior delivery outcome to indicate a subsequent pregnancy.(35) Records of early pregnancy loss that occur within 8 weeks of an early pregnancy loss outcome will be considered as linked to the same episode.

### Defining estimated date of conception

Several methods have been established for estimating date of conception using routinely collected data.(35–37) Based on these previous studies, we will derive the pregnancy start date using these variables in the following order of priority:

1. Estimated date of delivery (EDD)* minus 280 days

- We will prioritise EDD records that can be linked by date to an antenatal scan, as these are more likely to be accurate compared to EDDs recorded prior to the initial scan
2. Estimated date of conception (EDC)*
3. Date of last menstrual period (LMP)*
4. Date of in vitro fertilisation, intrauterine insemination or other assisted conception procedure (if applicable)
5. Antenatal scans – subtract week number (defined as number of completed weeks) of scan from date of scan (with preference towards earlier scans and those recorded in HES over GP records)
6. Recorded gestational age at delivery subtracted from delivery date
7. If none of the above are recorded, we will impute estimated date of conception as:

a. Date of full-term delivery minus 280 (252 to 287) days
b. Date of pre-term delivery minus 210 to 252 days
c. Date of multiple birth delivery minus 259 (238 to 273) days
d. Date of post-term delivery minus 287 days
e. Date of early pregnancy loss outcome (excluding ectopic pregnancy) minus 84 days (up to 180 days for miscarriage)
f. Date of ectopic pregnancy outcome minus 63 to 70 days

* Will only be used when date is not the same as the date of the record (I.e., the date that the GP input the estimated date into the system)

When dates are estimated or data considered less reliable, we will incorporate a degree of uncertainty in the estimation of the date of conception.

### Defining trimester start and end dates

The first trimester will be considered as the first 13 completed weeks (90 days) from the start of the pregnancy. The second trimester will be week 14 to week 26 (day 91 to day 181) and the third trimester will be week 27 (or day 182) until delivery.

## Statistical analysis plan

### Descriptive analysis

We will describe the overall pregnant cohort during the study period by the number of women with at least one pregnancy, average number of pregnancies for each woman in the cohort, demographic information, medical and pregnancy history and risk factors prior to first pregnancy. Risk factors include age at start of first pregnancy recorded in study period, ethnicity, deprivation, geographic region, smoking and alcohol intake, prior COVID-19 status, co-morbidities, including obesity (defined as BMI prior to pregnancy), diabetes, cardiovascular disease, mental health (e.g. depression), other factors associated with increased risk of severe COVID outcomes according to the QCovid algorithm, which is being used for vaccine prioritisation in the UK and recommendations from the Joint Committee on Vaccination and Immunisation. (24)

We will describe the number of women with missing antenatal scan records and other pregnancy records by ethnicity and deprivation. Additionally, we will describe the number of pregnancy and maternal outcomes by ethnicity and deprivation, including pre-eclampsia, gestational diabetes, caesarean or assisted birth, live single birth, live multiple birth, stillbirth (single/multiple), live and stillbirth for multiple pregnancy, pre-term birth, ectopic pregnancy, termination of pregnancy, miscarriage and other early pregnancy loss outcomes (molar pregnancy, blighted ovum).

We will describe the subset of data with mother-baby linkages by number of mothers, average number of linked babies per mother, year of birth, age, ethnicity, deprivation and geographic region of mothers and babies and smoking status, alcohol intake and co-morbidities of mothers. We will also describe the mothers who cannot be linked to baby records in the same way as above and babies who cannot be linked to mothers to report ethnicity, deprivation, age of mother at delivery (if recorded) and year of birth.

We will compare all three cohorts to determine whether there are significant differences between mothers and babies whose records can be linked versus those who cannot. We will use t-tests to compare continuous variables and chi-squared tests to compare categorical variables, where numbers are sufficient.

### A. Uptake of vaccines in pregnant women

#### Exposures

The exposures are date and type of first, second and third dose of COVID-19 vaccination, date of pertussis vaccination and date of influenza vaccination. We will include date of positive COVID-19 test result as a separate exposure.

#### Outcomes

The primary outcome is uptake of one or more COVID-19 vaccine dose in pregnant women. The secondary outcomes are uptake of second and third doses of COVID-19 vaccine and uptake of each type of COVID-19 vaccine for each dose. We will include Pfizer-BioNTech BNT162b2, Moderna mRNA-1273 and Oxford-AstraZeneca ChAdOx1-s/nCoV-19 vaccines in the analysis.

#### Descriptive analyses

We will tabulate overall uptake of each vaccine dose and vaccine type for women prior to and during pregnancy, by trimester and at delivery. We will also detail uptake of each dose of vaccine by population subgroups defined by age, ethnicity, deprivation, geographic region, smoking and alcohol intake, prior COVID-19 status, co-morbidities, prior anti-coagulant use and pregnancy history prior to study period.

For each vaccine type and dose, vaccination uptake over time (i.e., coverage by fetal age) will be estimated by the Kaplan-Meier method with pregnancy duration as the timescale. We will account for changes in vaccination policy over time by reporting outcomes separately in line with key policy change dates, particularly highlighting 16^th^ April 2021 as the date when all pregnant women were eligible for vaccination, rather than just those who were clinically vulnerable. We will also describe uptake by vaccine dose at different time periods relative to new COVID-19 variants, including the Delta and Omicron variants.

In addition to Kaplan-Meyer curves, we will report vaccine uptake by calendar month using graphs with dates of key policy changes highlighted.I]

#### Statistical analysis

We will calculate adjusted hazard ratios (95% CI) using Cox regression for uptake of each vaccine dose in pregnant women by pregnancy trimester and age group, ethnicity, deprivation, geographic region of the UK, smoking/alcohol intake, co-morbidities, and prior COVID-19 status. We will check for proportional hazards and extend to using Royston-Palmar models to account for time varying hazard ratios if the proportional hazards assumption is not valid. Analyses will be clustered at the GP clinic level where appropriate. Women will enter the analysis period on their pregnancy start date (or date on which vaccination is recommended if it is during pregnancy) and will be censored on the earliest date on which they deliver their baby, have a pregnancy loss, leave their practice, die or the latest date for which data are available.

##### Covariates in uptake analysis

Age, ethnicity, deprivation, geographic region, smoking and alcohol intake, prior COVID-19 status, co-morbidities, prior anti-coagulant use, pregnancy history prior to study period and previous vaccine dose given in pregnancy (for doses 2 and 3).

#### Sensitivity analyses

We will conduct several sensitivity analyses, including the exclusion of pregnancies that did not end in live birth, exclusion of time period before 16^th^ April 2021 and exclusion of pregnancies with inaccurate estimated date of delivery.

#### Secondary analysis

We will use identical methods to report uptake of influenza and pertussis vaccinations.

### B. COVID-19 vaccine effectiveness in pregnancy

#### Exposures

Vaccine exposure status during pregnancy will be treated as a time-varying exposure and defined as:I]

- Unvaccinated: no COVID-19 vaccinationI]I]
- Vaccinated: within 14 days from the first dose of vaccineI]
- One dose: from day 14 post first dose of vaccine to the earliest of end of pregnancy, date of second vaccine doseI]
- Two doses: from day 14 post second dose of vaccine to the earliest of end of study period, date of third vaccine doseI]I]
- Three doses: from day 14 post third dose of vaccine to the earliest of end of study periodI]I]
- Vaccinated before pregnancy onlyI]

We will include date of positive COVID-19 test result as a separate exposure and will identify women with SARS-CoV-2 infection or a combination of SARS-CoV-2 infection and vaccination during pregnancy.

#### Outcomes

The outcomes are COVID-19 hospitalisation, defined as hospitalisation with a code for COVID-19 or hospitalisation after a positive test for COVID-19 in the preceding 14 days (hospitalisations associated with a birth or pregnancy loss will not be considered an outcome) COVID-19 admission to intensive care, defined as admission to intensive care during a COVID-19 hospitalisation and maternal mortality from COVID-19, as defined by the MBRRACE-UK confidential enquiry into maternal deaths (Mothers and Babies: Reducing Risk through Audits and Confidential Enquiries across the UK).

#### Descriptive analyses

In the cohort of pregnant women, we will present numbers with COVID-19 outcomes by pregnancy trimester, vaccination status and subgroups defined by age, ethnicity, deprivation, geographic region, smoking and alcohol intake, prior COVID-19 status, co-morbidities, prior anti-coagulant use and pregnancy history prior to study period.

#### Statistical analysis

To investigate vaccine effectiveness, we will use incidence density sampling with replacement to match each case by age, calendar date, trimester of pregnancy and GP practice to controls without evidence of the outcome on that date, with a ratio of up to 1:10 cases to controls to match within the pregnant population. Individuals will be censored on the earliest of the following dates: date of outcome, date of death or study end date. Conditional logistic regression models will be used to estimate the odds ratios (ORs) for each COVID-19 outcome in the time periods described above following a first, second, third or fourth vaccine dose compared to unvaccinated people in the pregnant population. Vaccine effectiveness will be estimated as a percentage by calculating (1-OR)*100. We will conduct secondary analyses stratified by time periods when different SARS-CoV-2 variants were dominant: Alpha (18th December 2020 to 17th May 2021), Delta (18th May 2021 to 19th December 2021) and Omicron BA.1 (20th December 2021 to 1^st^ March 2022) (38).

##### Covariates in effectiveness analysis

Age, ethnicity, deprivation, geographic region, smoking and alcohol intake, prior COVID-19 status, co-morbidities, prior anti-coagulant use and pregnancy history prior to study period.

#### Sensitivity analyses

Sensitivity analyses include stratification by vaccination status prior to pregnancy, limiting COVID-19 hospitalisation to be defined as only those who had a positive PCR test within 14 days prior to admission, excluding pregnancies without accurate estimated date of delivery, excluding hospitalisations which are unrelated to COVID-19 such as postpartum haemorrhage and excluding hospitalisations related to a pregnancy event such as pre-eclampsia.

### C. COVID-19 vaccine safety in pregnancy

#### Study design

The primary analysis will be a cohort study. We will also conduct nested matched case-control studies, as they are an efficient design for the analysis of time-varying exposures. For each outcome we will identify cases (pregnant women or their babies depending on the outcome) with the outcome of interest and match up to 5 controls who did not have that outcome to each case by general practice, age of mother, calendar time and duration of pregnancy

#### Cohort study of maternal and perinatal outcomes

##### Exposures

Vaccine exposure status during pregnancy will be an aggregate categorical variable, defined by number of doses received during each trimester and prior to pregnancy. We will also incorporate type of vaccine for each dose.

##### Outcomes

Maternal outcomes include miscarriage, ectopic pregnancy, pre-term birth (iatrogenic, spontaneous or unknown; subdivided <28 weeks, 28-31 weeks, 32-36 weeks), pre-eclampsia, gestational diabetes and caesarean section or assisted delivery

Perinatal outcomesI]include stillbirth, early neonatal death, perinatal death, small for gestational age and congenital abnormalities. Congenital anomalies will be grouped according to EUROCAT guidelines as abdominal wall defects, central nervous system defects, circulatory system malformation, cleft palate, congenital heart defects, ear, face and neck anomalies, eye anomalies, gastro-intestinal anomalies, genital anomalies, kidney and urinary tract anomalies, limb anomalies, respiratory anomalies and genetic disorders. Each congenital anomaly will be classified as major or minor based on a blinded expert panel review.

##### Descriptive analysis

In the cohort of pregnant women, we will present numbers with safety outcomes by vaccination status and subgroups defined by age, ethnicity, deprivation, geographic region, smoking and alcohol intake, prior COVID-19 status, co-morbidities, prior anti-coagulant use and pregnancy history prior to study period. We will additionally describe the number of babies with recorded congenital anomalies and other perinatal outcomes in babies with linked mothers compared to babies without linked mothers, by age of mother (where recorded), ethnicity and deprivation.

##### Statistical analysis

We will use logistic regression analyses to determine unadjusted and adjusted odds ratios for the occurrence of each outcome by vaccination status (one, two or three doses; during each trimester and prior to pregnancy) compared to unvaccinated individuals.

Absolute and excess risks will be computed, combining information from the original cohort to estimate and compare the number of excess events following each type of vaccine (i.e. of the three COVID-19 vaccines used in the UK) and a SARS-CoV-2 infection.I]I]

Covariates: age, ethnicity, deprivation, geographic region, smoking and alcohol intake, prior COVID-19 status, co-morbidities, prior anti-coagulant use and pregnancy history prior to study period. For the analysis of congenital anomalies, we will additionally adjust for use of teratogenic medications immediately prior to and during pregnancy.

#### Cohort study of adverse events of special interest for vaccine safety

##### Exposures

Vaccine exposure status during pregnancy will be treated as a time-varying exposure and defined as:I]

- Unvaccinated: no COVID-19 vaccine
- Dose 1: monthly periods after first dose to the earliest of end of pregnancy or date of second vaccine dose
- Dose 2: monthly periods after second dose to the earliest of end of pregnancy or date of third vaccine dose
- Dose 3: monthly periods after third dose until end of study period
- We will also incorporate type of vaccine for each dose

For general vaccine safety outcomes occurring during pregnancy (i.e. not maternal or perinatal outcomes), we will only consider the risk period of 1 to 28 days after each dose as it is unlikely that outcomes occurring later than 28 days after vaccination would be related to the vaccine.

We will also include pregnancy trimester and SARS-CoV-2 infection as time-varying exposures.

##### Outcomes

Outcomes are adverse events in pregnant women of special interest for vaccine safety, including unplanned ICU admission, venous thromboembolism, myocarditis, myocardial infarction, arrhythmias, neurological outcomes such as Guillain Barre syndrome, ischaemic and haemorrhagic stroke and Bell’s palsy, blood disorders including idiopathic thrombocytopenia, aplastic anaemia and agranulocytosis and autonomic disturbances recorded as disorders of autonomic nervous system.

##### Descriptive analysis

In the cohort of pregnant women, we will present numbers with safety outcomes by vaccination status and subgroups defined by age, ethnicity, deprivation, geographic region, smoking and alcohol intake, prior COVID-19 status, co-morbidities, prior anti-coagulant use and pregnancy history prior to study period.

##### Statistical analysis

We will use time varying Royston-Palmar regression analyses to determine unadjusted and adjusted hazard ratios for the occurrence of each outcome by vaccination status (one, two or three doses) compared to unvaccinated individuals.I]Patients will enter the analyses on the estimated date of conception, calculated from birth date and gestational age at birth.I]I]Each woman will be classified according to whether they have been vaccinated or not, with vaccination as a time-varying exposure.I]Women will be censored on the earliest of date of outcome of interest, death, end of pregnancy or end of the study period or last date for which data are available at the time of the analysis.I]

Covariates: age, ethnicity, deprivation, geographic region, smoking and alcohol intake, prior COVID-19 status, co-morbidities, prior anti-coagulant use and pregnancy history prior to study period.

Absolute and excess risks will be computed, combining information from the original cohort to estimate and compare the number of excess events following each type of vaccine (i.e. of the three COVID-19 vaccines used in the UK) and a SARS-CoV-2 infection.I]I]

#### Matched nested case-control studies

##### Selection of cases and matched controls

We will identify cases of each outcome (pregnant women or their babies depending on the outcome) and match up to 5 controls without that outcome to each case by general practice, age of mother, calendar time and duration of pregnancy. I]

##### Exposures

Vaccine exposure status during pregnancy will be an aggregate categorical variable, defined by number of doses received during each trimester and prior to pregnancy. We will also incorporate type of vaccine for each dose.

##### Statistical analysis

Conditional logistic regression analysis will be used with case and control matching groups as the strata. Adjustments will be made separately for confounding factors associated with the increased risk of each outcome and together in the maximally adjusted model.I] Odds ratios will be calculated for vaccinated versus unvaccinated women. Unadjusted and adjusted odds ratios, along with 95% CIs, will be calculated.I]We will adjust for the same covariates as in the cohort study.

Absolute and excess risks will be computed, combining information from the original cohort to estimate and compare the number of excess events following each type of vaccine (the three COVID-19 vaccines used in the UK) and a SARS-CoV-2 infection.I]I]

#### Secondary Analyses

Identical methods will be used to evaluate safety of influenza and pertussis vaccination in pregnant women.I]

##### Sample size calculations

The number of live births in England and Wales in 2020 was 615,557 (33). As a conservative estimate of the number of pregnancies, we estimate that there were 500,000 pregnancies in England in 2020. QResearch consists of a 20% sample of the general population, meaning we are likely to have approximately 100,000 pregnancies in 2021 and 40,000 up to May 2022 giving in excess of 140,000 for inclusion in the cohort studies (objectives A and B).

#### Vaccine effectiveness sample size (Objective B)

The prevalence of the severe COVID-19 outcomes in the general population is less than 3%. (24) Using the prevalence of the outcomes in the general population we computed the minimum sample size needed to estimate a conservative hazard ratio of 0.8 (corresponding to 20% vaccine effectiveness) or 0.5 (corresponding to 50% vaccine effectiveness) for developing one of the primary endpoints, with significance of 0.05 and power of 0.8. We anticipate that we would need at least 631 pregnant women with each outcome and a cohort of at least 63,053 pregnant women to be able to detect an HR of 0.8 with significance at 0.05 and power of 0.8 for the less prevalent outcome (0.01 event probability).

| HR | Vaccine effectiveness | Event probability | Number of cases | Sample size |
| --- | --- | --- | --- | --- |
| 0.8 | 20% | 0.03 | 631 | 21,018 |
| 0.8 | 20% | 0.02 | 631 | 31,527 |
| 0.8 | 20% | 0.01 | 631 | 63,053 |
| 0.5 | 50% | 0.03 | 66 | 2,179 |
| 0.5 | 50% | 0.02 | 66 | 3,268 |
| 0.5 | 50% | 0.01 | 66 | 6,535 |

#### Vaccine safety sample size (Objective C)

We assume different exposure probabilities in the control group (patients without adverse pregnancy outcome) and estimate the number of cases (patients with each adverse pregnancy outcome) required to detect an OR of 1.2 with significance level of 0.05 and power of 0.8, considering a matching ratio of 5 controls per case.

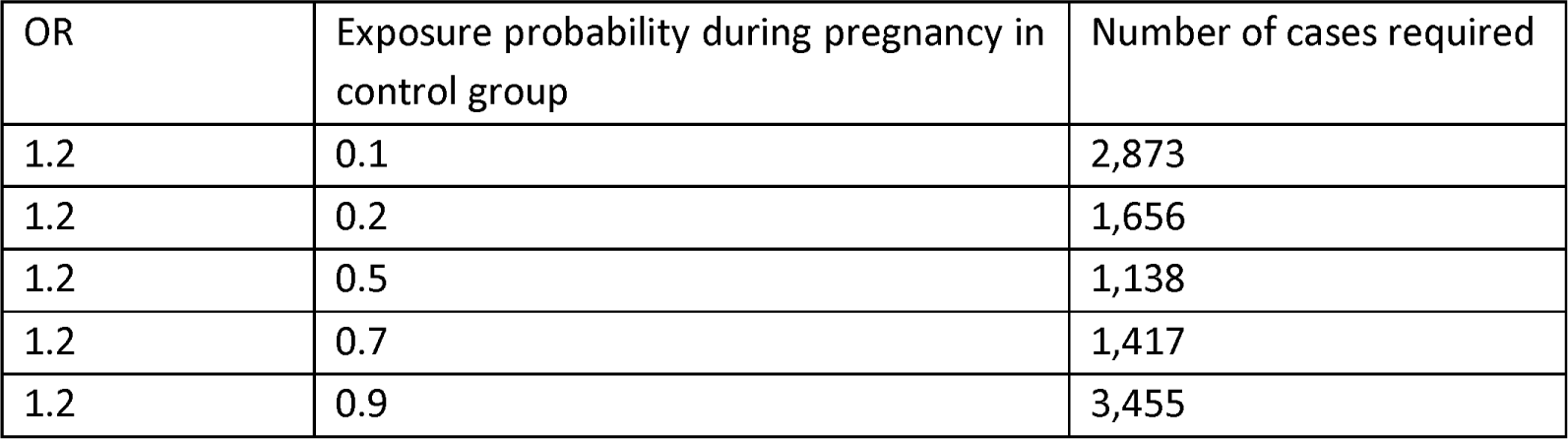

| OR | Exposure probability during pregnancy in control group | Number of cases required |
| --- | --- | --- |
| 1.2 | 0.1 | 2,873 |
| 1.2 | 0.2 | 1,656 |
| 1.2 | 0.5 | 1,138 |
| 1.2 | 0.7 | 1,417 |
| 1.2 | 0.9 | 3,455 |

##### Plans for addressing missing data

For all analyses, we will initially conduct complete case investigations. We will subsequently evaluate the models in multiply imputed data. Under the ‘missing at random assumption’, we will use multiple imputation with chained equations to generate 5 imputed datasets, where values for ethnicity, body mass index (BMI), Townsend deprivation quintile and smoking status are imputed.(39–42) Imputation models will include all exposure and outcome variables; statistical models will be developed on each of the 5 imputed datasets and estimates pooled using Rubin’s rules.

##### Strengths and limitations

This study has several strengths. It will be the largest analysis examining COVID-19 vaccination outcomes in pregnancy. Setting up linkages will enable a definitive analysis of the associations between vaccination and safety outcomes in pregnancy including rare outcomes such as congenital anomalies. Establishing the linkages and methodology will facilitate future work to investigate post-marketing vaccine safety and effectiveness in pregnant women. Furthermore, linkages between mothers and babies will allow a future analysis into longer-term health and development outcomes for the children. In addition to reporting outcomes from COVID-19 vaccination this work will report outcomes from more established vaccinations commonly used in pregnancy (influenza and pertussis) which will help clinicians communicate risks and benefits of vaccination in pregnancy.

Limitations of our study include potential lack of formal adjudication of diagnoses (e.g., comorbidities on Read codes and failure to register lateral flow test results), potential for misclassification of outcomes, information bias and potential bias due to missing data. However, since we have complete national datasets for exposures (such as the NIMS vaccination status) and outcomes (such as critical care and mortality), we anticipate minimal impact on the robustness of our findings. It can be very difficult to determine pregnancy trimester using GP data alone since not all the information is consistently recorded. However, we anticipate that the use of the already linked HES records and the additional MSDS will enable us to develop an algorithm to estimate this based on primary care data plus gestational age at delivery and delivery date. Additionally, we will carry out sensitivity analyses when uncertainty in date of conception remains. Our study is not randomised and so has less utility for determining vaccine effectiveness compared with a conventional trial. However, in the absence of very large-scale post marketing randomised trials particularly in pregnant women observational assessment of the risk of COVID-19 diagnoses and outcomes of COVID-19 infection following vaccination will provide the best available information, albeit with a cautious interpretation. We have already begun drawing up data sharing agreements to establish linkages between QResearch and new datasets, NCARDRS Congenital Anomaly register and Maternity Services data, but there remains a possibility that these cannot be completed within the time frame of the study which may limit some of the outcomes that we are able to assess for some women.

Some variables and outcomes, including miscarriage, will not be well characterised in the dataset meaning we will have incomplete data. We will only have access to PCR test data for positive COVID-19 test results. This will result in an underestimation of the number of positive COVID-19 cases as some people will have tested positive using a lateral flow test and others will have contracted COVID-19 prior to the widespread roll-out of testing. This could potentially result in an underestimation of vaccine effectiveness.

#### Patient and public involvement

As a project team, we have established relationships with parallel PPIE panels. These PPIE panels include participants from the Centre for Ethnic Health Research and participants with lived experience of pregnancy and vaccination.

These have together helped us develop our proposal. They tell us that understanding more about uptake and safety of COVID-19 vaccination pregnant women remain important research questions. We have spoken with pregnant women who are considering COVID vaccination. They told us how they are significantly concerned about what they identify as a lack of evidence about COVID vaccination in pregnancy. These include risks to them, but also risks to the pregnancy (for example, impacts on fertility, risk of miscarriage, risk to the baby’s growth leading to intra-uterine growth restriction or pre-term labour) and risks to the baby (including developmental defects present at birth and longer-term risks). Women with experience of making decisions about whether or not to have a COVID vaccination in pregnancy have told us that they found this difficult, and that wanted more information about a wider range of risks, both to themselves, the baby, and the pregnancy. They agreed that knowing that the risks of COVID were significant was important, but they wanted more detail and more information. They told us that the information this study will look for would have been helpful and of value for them. This included having more information to share with family members at home who were also concerned about vaccine safety or necessity in pregnancy. If possible, they would like the information to be tailored, so that women could make individualised decisions – for example including the context of ethnicity or other long-term conditions. Finally, the timing of the vaccines in pregnancy, and whether this affects risks was a question they wanted more information about. The safety of the vaccines (compared against COVID) in the context of their illnesses and treatment regimens are questions of paramount importance to them are supportive of this research question and approach. They value a research approach which can be responsive to emerging concerns about vaccination and COVID. They advised us that exploring the impacts of the COVID booster programme, including where the booster vaccine is different from the vaccine used in the primary schedule is vital to explore. They support using the surveillance methods proposed in this project (having worked with us and developed an understanding of this approach over the last year).

Finally, they told us that understanding the long-term effects for a wide range of people was an important research ambition, highlighting a need to continue monitoring and developing our understanding of the impacts of vaccination through the pandemic.

We have established integrated PPIE support from both community settings (supported by the NIHR’s Applied Research Collaborative East Midlands, ARCEM) and the pregnancy community via the Maternity Voices Partnership and through Dr Brenda Kelly (England) and Prof Aziz Sheikh (Scotland). Their guidance has been integral to our previous research work design and development. For example, they advised us to contextualise vaccine risk against the risks of COVID infection and have strongly supported the development of research that furthers understanding of vaccine risks for those with any long-term conditions. They are worried about the lack of information about the COVID-19 vaccines in pregnant women and strongly support us gathering evidence to support women, families and policy makers in their decision making. As with our previous vaccine research, we will ensure that we have PPIE representation from diverse communities and from the groups who are at the heart of our study. For this study, we will develop a PPIE group including women with experience of making decisions about COVID-19 vaccination whilst pregnant. The ARCEM team have established PPIE groups which include women with experience of decision making about COVID-19 vaccination in pregnancy, and this will therefore be accessible to inform the study from the beginning.

Ongoing input throughout the research cycle will help us ensure that our research remains important to the wider community as boosters are given. With our PPI team we will co-create publications for a lay audience, to make sure these are clear, culturally adapted, accessible, interesting and informative. This will ensure that pregnant women will be supported to make an evidence-based informed choice about vaccines uptake. The panel may provide insights and may help people understand how the research relates to them. We will work with diverse PPI advisers to support the communication our findings. We will continue to offer bespoke training and support.

Research findings will be submitted to pre-print servers such as MedRxIv, academic publication and disseminated more broadly through media releases and community groups and conference presentations. Panel members will be invited to suggest media to reach important audiences, for example media that caters for diversity. We will continue to co-design easy-to-understand infographics in collaboration with patients, as we did for the risk of blood clots associated with different types of COVID-19 vaccination.(30)

#### Study Management

Six management meetings are planned over the duration of the study, including all co-applicants, collaborators and PPI representatives:

1. Oct 2022 - Identification of important outcomes and study population
2. Apr 2023 – Update of study progress. Description of study cohort by ethnicity, geography and socioeconomic status
3. July 2023 – Update on study progress - Results on vaccine uptake, overall, by trimester of pregnancy and by ethnicity, geography and socioeconomic status
4. Dec 2023 - Update on study progress - Results on vaccine effectiveness, overall, by trimester of pregnancy and by ethnicity, geography and socioeconomic status
5. May 2024 - Update on study progress - Results on vaccine safety, overall, by trimester of pregnancy and by ethnicity, geography and socioeconomic status
6. Sept 2024 – Final results and dissemination plan

## Data Availability

All data produced in the present work are contained in the manuscript

